# Droplet digital PCR assay to analyze allele-specific mRNA expression on HTT repeat expansion locus

**DOI:** 10.1101/2025.09.26.25336715

**Authors:** Eugenio Gentile, Michel Tessier, Jessica Migliavacca, Andrea Manfrin, Nazia Maroof, David J Hawellek, Marc Sultan, Damian Roqueiro, Anna Rautanen

## Abstract

**Background:** Huntington’s disease (HD) is a fatal neurodegenerative disorder caused by a mutation in the huntingtin gene (HTT), characterized by an expanded CAG trinucleotide repeat. At the time of writing no cure or disease-modifying treatments exist. Currently, the most explored investigational therapeutic strategy targets HTT gene expression, either lowering both alleles (total lowering) or selectively lowering the mutant allele. These approaches require reliable pharmacodynamic biomarkers to measure an allele-selective knockdown. However, allele specific quantification of wild-type and mutant HTT RNA or protein remains a challenge.

**Results:** Here we optimized a droplet digital PCR (ddPCR) assay to distinguish between mutant and wild-type HTT (mHTT and wtHTT respectively) mRNA expression based on differential amplification of HTT mRNA molecules with different CAG repeat lengths under limited dNTP conditions. This assay, combined with our novel automated analysis pipeline reliably detects allele-specific expression in HD patient cell lines. We simulated various mHTT to wtHTT mRNA ratios by mixing RNA from respective homozygous cell lines to demonstrate the assay’s accuracy under varying allele ratios. We also validated the assay’s utility in 13 cell lines from HD patients and their family members. Additionally, we optimized a one-step RT-ddPCR method, offering a streamlined alternative to a two-step ddPCR method. We further confirmed the assay’s clinical relevance by demonstrating allele-selective siRNA mediated HTT lowering in HD patient fibroblast cell lines.

**Conclusions:** Our optimized ddPCR assay, with its pipeline for automated data analysis, enables the precise quantification of allele-selective HTT mRNA knockdown. The method does not require prior knowledge of patients’ SNP genotypes, previously a prerequisite for assays aiming to determine the mHTT transcript expression in patient samples. Our HTT ddPCR assay is universally applicable regardless of patient genotype. The ability to accurately monitor allele-specific HTT mRNA expression levels holds great promise for developing effective treatments for Huntington’s disease.

## Background

Huntington’s disease (HD) is a fatal neurodegenerative monogenic disorder that is caused by a mutation with an autosomal dominant inheritance in the huntingtin gene (HTT) on chromosome 4 (1). The mutation involves a somatically unstable, expanded CAG trinucleotide repeat in exon 1 of HTT that translates into a protein with an expanded polyglutamine tract (2). The resulting underlying molecular pathogenesis of HD is complex and the precise mechanism of toxicity is still debated. It is believed that toxicity is primarily driven by misfolding of the mutant HTT (mHTT) protein, which then accumulates in the brain, leading to neuronal dysfunction and cell death (3). Additionally, the somatic instability of the CAG repeats in the HTT gene itself is thought to play an important role (2,4). The number of CAG repeats varies between individuals and correlates with the onset and severity of the disease (5,6). The length of the CAG repeat is considered normal (wild-type), when the number of repeats is less than 36 and pathogenic (mutant) when the number is 40 or more (7). Individuals with a number of repeats between 36 and 39 may or may not develop HD, i.e. they have reduced penetrance (8). Symptomatically, HD is characterized by a triad of motor, cognitive, and psychiatric dysfunction, manifesting as a progressive decline in voluntary movement control, intellectual abilities, and emotional stability, ultimately leading to death (9). Currently, HD has no cure or available treatments to slow or stop the progression of disease (4,10). One of the most attractive strategies under development for the reduction of the quantity of mHTT protein in the brain are antisense oligonucleotides (ASOs), siRNA and miRNA approaches that aim to specifically target the mHTT transcript while leaving the wild-type transcript (wtHTT) free to be translated (11). These treatment approaches however require purpose-built pharmacodynamic biomarker assays to demonstrate the HTT allele-selective knockdown on the mRNA level.

In order to address this gap in the field, here we have further developed an allele-specific HTT mRNA ddPCR assay that was originally described by Dodd et al. in 2013 (12). The described assay relies on ddPCR conditions with limited dNTP availability to differentiate between mHTT and wtHTT transcripts. A restricted dNTP availability in the ddPCR mix indirectly limits the number of amplification cycles, allowing the shorter wtHTT transcript (with fewer CAG repeats) to amplify more than the longer mHTT transcript before the available dNTPs are depleted. This results in a stronger fluorescence signal for the droplets containing the wtHTT transcript, creating two distinct clusters (Fig. 1a). In ddPCR, the input sample is partitioned into approximately 20,000 individual droplets where each positive droplet contains at most one PCR target molecule. Each droplet then undergoes its own, independent PCR reaction as part of a suspension. The number of “positive” droplets, containing the target transcript, determines the absolute concentration of the mRNA sequence targeted by the PCR primers in the reaction mixture. In our case, the fluorescence amplitude of each droplet detected by the ddPCR droplet reader (the intensity of the fluorescence signal), is used to differentiate between mHTT and wtHTT transcript-containing droplets (Fig. 1). In contrast, conventional PCR assays achieve allele-specific detection by designing primers that exploit nucleotide level differences between the different HTT alleles. However, this allele-specific primer design philosophy cannot be directly applied to nucleotide repeat regions. Recently published studies discriminate between mHTT and wtHTT transcripts based on Single-nucleotide polymorphisms (SNPs) that are in high linkage-disequilibrium (LD) with the CAG repeat region (13–15). However, measuring allele-specific HTT expression reliably via SNPs would require long-read sequencing or other advanced methods that are able to phase the CAG repeat with the SNP of interest. This is technically extremely challenging, and therefore previous studies describing allele-specific HTT mRNA detection are purely relying on published, population level LD between those two genetic loci. This approach, however, is prone to imprecisions as the level of LD varies significantly between different populations and, moreover, requires heterozygosity of those reference SNPs in the analyzed patient samples. In contrast, the method described here does not rely on specific genotype composition, making it suitable for determining the mHTT concentration within a wider range of patient samples.

**Fig. 1.**
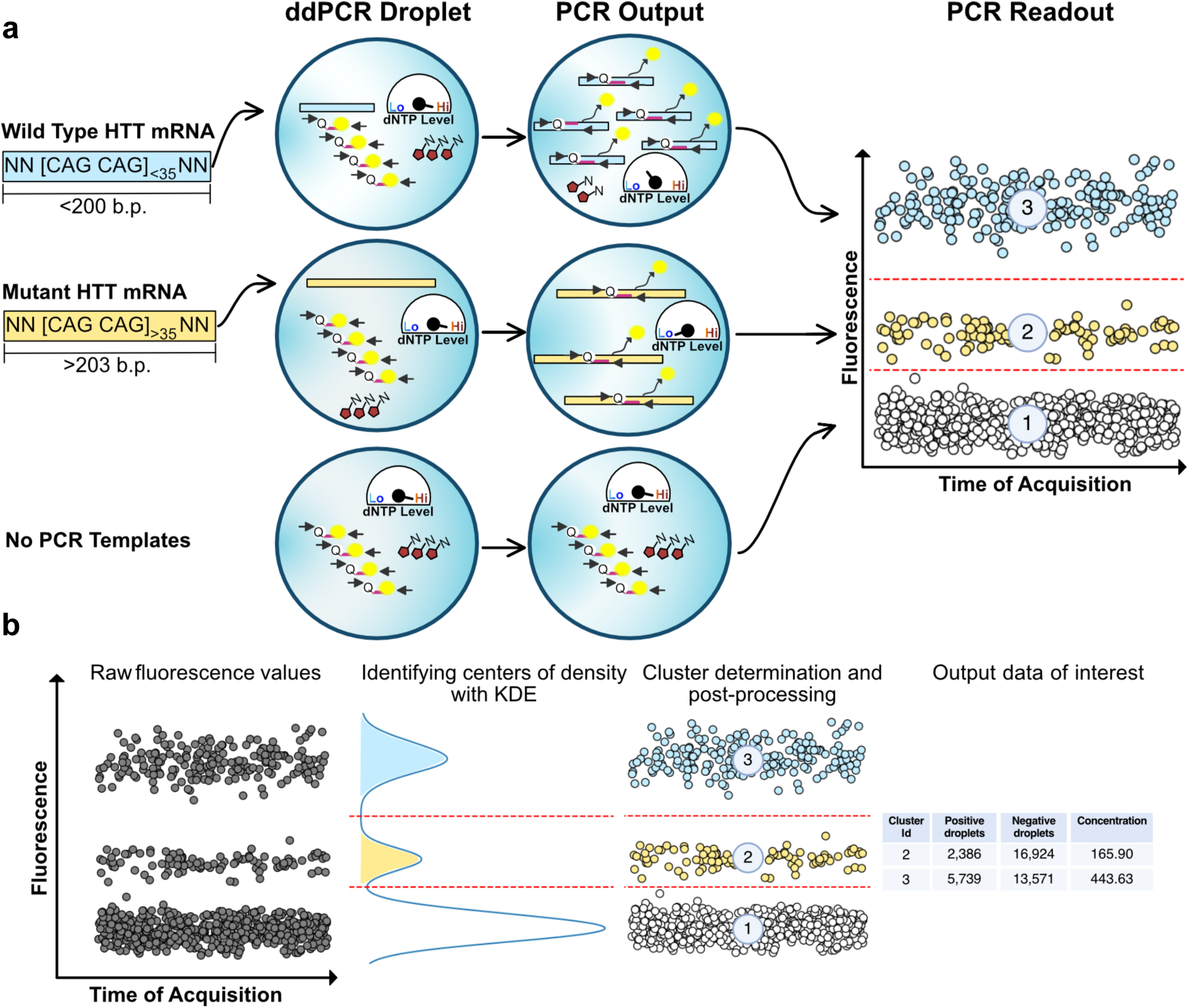
Principle of the allele-specific HTT ddPCR assay and the developed bioinformatic data analysis pipeline. **a)** The assay is based on ddPCR conditions in which the availability of dNTPs (marked by red hexagons and a gauge showing the amount of remaining dNTPs) is a limiting factor. Therefore, the shorter wild-type repeat (fragment marked in light blue) can go through more rounds of amplification than the longer mutant repeat (fragment marked in light yellow) before the dNTPs are depleted. In ddPCR, the sample is partitioned into approximately 20,000 individual droplets (1 droplet per scenario is illustrated in this figure), and each droplet undergoes an independent PCR reaction. As a result of the dNTP depleted conditions, the fluorescent signal is stronger for the wild-type HTT transcript-containing droplets than for the mutant allele-containing ones, and two distinct positive clusters are created (cluster 2 represents the mutant allele and cluster 3 the wild-type allele), in addition to the negative (empty) droplet cluster (cluster 1). As in standard ddPCR, the number of positive droplets is counted to quantify the absolute concentration of the mRNA. Droplet fluorescence amplitude (the intensity of the fluorescence signal) is in our case utilized only for differentiating the mutant and wild-type clusters, rather than influencing the actual quantification. **b)** The droplets and their raw fluorescence values are shown in the left panel. Conceptually, to determine the thresholds that separate the clusters, we can rotate the plot and have the fluorescence values as the X-axis. If we then compute the density on these fluorescence values, we will obtain a density curve with 3 peaks, corresponding to the 3 centers of mass of the clusters (1. Negative droplets; 2. Positive droplets with longer amplicon; 3. Positive droplet with shorter amplicon). The thresholds (shown as red dotted lines) are then obtained as the minimum values of each of the valleys between the peaks. With these thresholds, the droplets are assigned to clusters and the concentration of each cluster in the experiment is computed.

The aim of this study was to advance this unique assay principle and to prove its wide applicability by testing it using a range of biological samples. To achieve this, we first optimized the protocol for accurate measurements of mutant and wild-type HTT mRNA and tested the reproducibility and accuracy with several different healthy and HD patient-derived samples. Following assay optimization, we further demonstrated the assay’s utility for biomarker detection purposes during drug development by assessing allele-selective HTT mRNA knock-down following allele-selective siRNA treatment. Importantly, we also developed an automated analysis pipeline for objective and reproducible identification and quantification of wtHTT and mHTT transcript droplet clusters within the ddPCR data obtained from our experiments.

## Methods

### Sample description

Altogether 13 Human Fibroblast cell lines from HD patients and their family members were used in this study with varying CAG repeat lengths and differing homozygous/heterozygous statuses (Supplementary Table 1). Cell lines were obtained from NIGMS Human Genetic Cell Repository at the Coriell Institute for Medical Research, Camden, New Jersey.

### Cell culture conditions

All 13 fibroblast cell lines used for experiments were maintained in high glucose Dulbecco’s Modified Eagle Medium (DMEM, Gibco, #10313021) containing 10% fetal bovine serum (FBS, Gibco, 26140079), 1% MEM non-essential amino acids (NEAA; Gibco, #11140050), 1% GlutaMAX (Gibco, #35050061), and 1% penicillin/streptomycin (Gibco, #15140122), and incubated at 37 °C with 5% CO_2_.

### Total RNA extraction

Total RNA was extracted from approximately 2 × 10^5^ fibroblasts using the RNeasy Extraction Mini kit (Qiagen, #374104) following the manufacturer instructions and protocol including the use of DNase treatment. High RNA quality was confirmed using an Agilent 2100 bioanalyzer (Agilent Technologies).

### Droplet Digital PCR (ddPCR)

Both 2-step and further optimized 1-step RT-ddPCR approaches were used in this study. ddPCR reaction droplets were generated using a Bio-Rad Auto Droplet Generator (AutoDG) instrument (BioRad), following the manufacturer’s instructions.

In the 2-step RT-ddPCR approach, 200 ng of total RNA was used for cDNA conversion in a final volume of 20 μL using SuperScript IV VILO Master Mix with ezDNase Enzyme according to the manufacturer’s protocol (Thermo Fisher, #11766050). A volume of 2 μL of the undiluted cDNA was amplified using primer/probe mixture in 1.5 µM and 0.5 µM final concentrations, respectively, with 1x ddPCR mastermix (Supermix for Probes) (BioRad, #1863026). The used primer/probe concentration is higher than in standard ddPCR to create dNTP limiting conditions. Primer and probe sequences have been previously described by Dodd et al. (12) (Forward primer: GGCTGAGGAAGCTGAGGAG; Reverse primer: ATGGCGACCCTGGAAAAG; Probe: TTCGAGTCCCTCAAGTCCTTCC with 6-FAM dye and 36/TAMRA Sp modification). For experiments using GUSB as a housekeeping gene, Bio-Rad Assay ID dHsaCPE5050189 was used in standard conditions (900 nM/250 nM primer/probe final concentration). Thermal cycling conditions were the following: denaturation at 95°C for 10 min followed by 40 cycles of 94°C for 30 sec and 60°C for 30 sec, followed by enzyme inactivation at 98°C for 10 min and cooling to 4°C for at least 15 min before further processing. After amplification, droplets were analyzed on a QX-200 ddPCR Droplet Reader instrument (BioRad), following manufacturer’s instructions.

For the 1-step RT-ddPCR approach, 50 ng of RNA were used as starting material together with the One-Step RT-ddPCR Advanced Kit for Probes (Bio-Rad, #1864022) according to the manufacturer’s protocol. The cycling conditions were identical as above except for an additional step corresponding to the reverse transcription at 50°C for 1 hour before PCR amplification.

### Simulation of the allele-selective lowering

cDNA from mHTT (GM13511) and wtHTT (GM04859) homozygous cell lines were mixed in predefined proportions (mHTT:wtHTT ratios of 100:0, 80:20, 60:40, 50:50, 40:60, 20:80 and 0:100) in 3 technical replicates to simulate the effect of allele-specific lowering. To measure the mHTT vs. wtHTT allele proportion, ddPCR reactions were performed using a 2-step approach as described above.

### siRNA transfection

The HD patient fibroblast cell lines (Coriell GM04022 and GM02147) were transfected at ∼60% confluence with 10 nM or 100 nM siRNAs using Lipofectamine RNAiMAX Transfection Reagent (Thermo Fisher Scientific, #13778075) following the manufacturer’s instructions. Two different allele-specific siRNAs targeting either the cytosine (5’-GAA<u>G</u>UACUGUCCCCAUCUC-dTdT-3’) (16) or the adenine (5’-GAA<u>T</u>UACUGUCCCCAUCUC-dTdT-3’) at the SNP position rs363125 were used. Non-targeting Control siRNA (Qiagen, #1022076) was used as a negative control for siRNA transfection. Total RNA from two biological replicates was extracted 72h after transfection. A 2-step RT-ddPCR approach was then performed to determine the allele-specific changes in HTT expression following treatment with SNP-selective siRNAs.

### Data analysis

To avoid the manual setting of thresholds, a pipeline to determine the optimal thresholds between clusters of ddPCR droplets was developed in-house. ddPCR droplet fluorescence amplitude readouts were exported using Biorad’s QX Manager Software 2.0 (17) as comma-separated files (.csv). The .csv files were used as inputs for an in-house-developed analysis pipeline to perform our automatic threshold detection (Fig. 1).

Briefly, we separate the raw fluorescence values of the detected droplets into clusters by estimating the density function of the fluorescence values (Fig. 1b). Computing the density on the fluorescence values will yield a number of peaks that corresponds to the center of mass of the identified clusters. The boundaries, i.e., thresholds, between these clusters are then obtained by finding the minimum value of the valleys between the peaks.

The thresholds obtained with our method avoid manual setting and bring reproducibility to the entire downstream analysis process. The concentration for each cluster of droplets was computed using a modified formula based on one provided by the manufacturer (described in the “concentration calculation” section below).

The results presented in this paper are based on our automatic pipeline, unless stated otherwise. The pipeline contains four main modules, which are detailed below.

### Preprocessing

To detect outliers we implemented the local outlier factor (LOF) algorithm (18). The LOF algorithm is an unsupervised method to perform anomaly detection that flags a data point as an outlier if its value deviates from a local density measure computed among the neighbors of the data point. The implementation of LOF was done using Sci-kit learn (19) with a parameter of number of neighbors set to 20. Droplets with amplitude values that were considered outliers were removed from the analysis. An example of the outlier removal process is shown in Supplementary Fig. 1.

### Cluster identification

The distribution of amplitude values was used as input to the process of cluster identification. To automatically detect the boundaries between clusters, a univariate version of the Kernel Density Estimation (KDE) was used (20). KDE is an unsupervised method to estimate the density function of a distribution of values. The KDE is used as a proxy to the true density of amplitude values. Alternative approaches to density estimation like binning of data followed by the computation of histograms did not provide the resolution needed to identify the exact boundaries between clusters. KDE was implemented using Sci-kit learn (19). A key parameter in this method is the smoothing factor *h*, also known as bandwidth. A cross-validation scheme with a grid search to explore the parameter space for *h* was used (values of *h* between 10 and 150, in increments of 10 were used in the search). The value of *h* = 100 used for this analysis was obtained as a tradeoff between i) the optimization of *h* for KDE and ii) the smoothness of the KDE curve for peak and valley detection as detailed in the next paragraph. The dataset “Simulation of the allele specific lowering”, described in the results section, was used to fine-tune the bandwidth. In this dataset, the ground truth assignment of droplets to clusters is known.

Analysis of the KDE was performed to find the peaks in the curve. Each peak corresponds to the center of mass in a cluster. The thresholds between clusters were obtained by finding the minimum value in the valleys between two peaks (Fig. 1b).

### Post-processing

A post-processing step was performed in each of the detected clusters. Clusters with an extremely low density of droplets (less than 0.05% of total droplets) were merged with the cluster whose centroid was nearest to the centroid of the cluster to remove. This was done to avoid reporting clusters that represent noise (often described as “rain”) rather than a real biological signal. If not removed, these spurious clusters would artificially create an additional “allele cluster”. By setting the 0.05% droplet clustering threshold, we also set the lower limit of detection.

### Computation of concentration

In an experiment with *k* clusters, for cluster *i* with *i* ∈ {1, 2, … *k*}, the concentration is computed as:

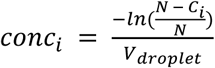

where *N* is the total number of droplets, *C*_*i*_ is the number of droplets in cluster *i* and *V*_*droplet*_ is the average volume of the individual ddPCR droplets (calculated by the QX200 droplet reader).

## Results

### Analysis pipeline

The default ddPCR data analysis software, QX Manager (Bio-Rad), allows users to manually set the threshold between the high (wtHTT) and low (mHTT) amplitude clusters to calculate the detected copies per microliter of the assay target in the given clusters. However, this manual process is prone to human error and suffers from inter-user variability. To avoid this manual setting of thresholds, and to maximize reproducibility, an in-house developed computational pipeline was used to analyse the HTT ddPCR data obtained (Fig. 1b). Within our pipeline we included an outlier removal step to prevent droplets with anomalous fluorescence amplitudes from skewing the data (Supplementary Fig. 1). In general, the number of outliers was low (0.1% on average per well, data not shown). In the cluster identification step we obtained the thresholds separating the clusters for each well. These thresholds were then used to compute the concentrations of all clusters.

### Simulation of HTT allele specific lowering

To evaluate the accuracy of the assay’s ability to detect the correct mutant to wild-type HTT allele ratio, we mixed isolated total RNA from mHTT and wtHTT homozygous cell lines. This created a series of samples ranging from complete homozygosity for the wild-type allele (GM04859 with a 20/20 CAG repeat HTT genotype) to complete homozygosity for the mutant allele (GM13511 with a 47/45 CAG repeat HTT genotype). These mixtures can be seen as a simulation of a differential expression of the two alleles, mimicking drug-related allele-selective lowering. Across the varying wtHTT:mHTT allele ratios, we observed minimal technical variability between the three technical replicates, and the reduction in mHTT proportion matched the expected (input) proportions of the mHTT allele (Fig. 2a, b, and Supplementary Table 2). Therefore, these results confirm both the high accuracy and reproducibility of our assay, as evidenced by the strong correlation between expected and detected mHTT percentage and the minimal technical error, respectively (Fig. 2b).

**Fig. 2.**
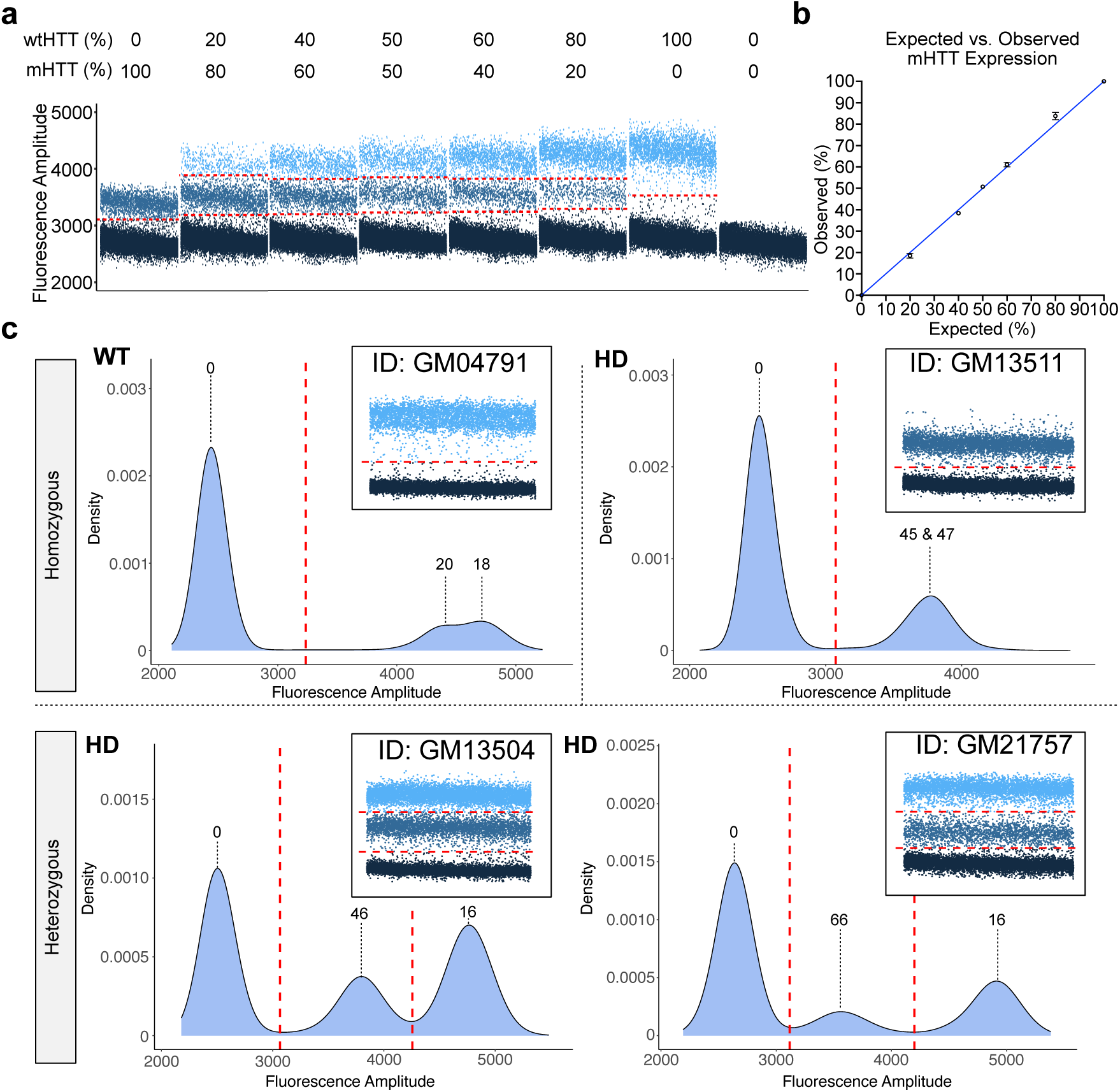
Simulation of allele-specific lowering and examples of allelic separation in different patient-derived cell lines. **a)** Simulation of the allele-specific lowering by mixing mHTT (GM13511; 45/47 CAG repeats) and wtHTT (GM04859; 20/20 CAG repeats) homozygotes in known proportions. Distribution of the droplets in pre-defined mHTT proportions: 100%; 80%; 60%; 50%; 40%; 20%; 0%; no input control. **b)** Scatter Plot with standard deviation error bars shows the observed vs expected mHTT expression level proportions in 3 technical replicates for each mHTT proportion tested. **c)** Examples of allelic separation of HTT CAG repeat region in ddPCR in selected Coriell cell lines (GM04791, GM13511, GM13504, GM21757). The negative droplet cluster is marked as 0, whereas the positive droplet clusters are marked based on the known CAG repeat lengths (determined using sequencing).

### HTT Allele-specific mRNA expression of 11 Coriell samples

In order to assess the utility of the assay in variable CAG repeat lengths typical for Huntington’s disease patients, we measured allele-specific mRNA expression of 11 different Coriell samples (Supplementary Table 1) from Huntington’s disease families including both patients and family members. Our results show that two different alleles (e.g. different lengths of the CAG repeats) produce two distinct clusters when amplified in dNTP depleted conditions (Fig. 2c, Supplementary Fig. 2, and Supplementary Table 1). As small as a 10% difference in the repeat length could be discerned visually (as demonstrated by the Coriell sample GM04791 with 18/20 repeat length units), although the clusters were still largely overlapping (Supplementary Fig. 2). To improve the sensitivity of our cluster detection pipeline and assess whether allele clusters could be better resolved in GM04791, we applied a lower bandwidth factor (*h* = 90). This adjustment enabled reliable identification of a threshold separating the two wtHTT alleles, allowing us to quantify each allele’s expression individually (Supplementary Fig. 3). For comparison, the difference between mHTT homozygous alleles of GM13511 with 45 and 47 CAG repeat length units (a 4.4% difference in length) can no longer be differentiated visually or with our automatic cluster detection pipeline, regardless of bandwidth lowering (Supplementary Fig. 2 and Supplementary Fig. 3). Throughout all tested heterozygous samples the mHTT and wtHTT droplet clusters are clearly separated from each other (Fig. 2c and Supplementary Fig. 2). Additionally, we observed that the transcripts with higher CAG repeat numbers consistently showed lower expression levels compared to the transcripts with lower CAG repeat numbers (Supplementary Fig. 2). This cannot be explained by differential amplification efficiency between the different lengths due to the absolute quantification nature of ddPCR - differential amplification efficiency is utilized in this approach only to separate the mHTT and wtHTT clusters.

To investigate the effect of cDNA input on HTT transcript detection, we performed 5-fold cDNA dilutions of the Coriell samples (Supplementary Fig. 4, full data not shown). While the total number of positive droplets decreased with dilution, PCR amplification efficiency slightly increased per positive droplet. This improved amplification efficiency, as shown in the figure, is particularly beneficial for longer CAG repeat lengths, as it enhances the separation of their clusters from the negative droplet cluster, reducing overlap and improving differentiation).

To optimize the RT-ddPCR further, we analyzed a subset of the same Coriell samples (n=8) using 1-step RT-ddPCR approach (2-step ddPCR approach was described by the original paper (12)) and automated thresholding and concentration calculations instead of Biorad’s QX Manager software. Results were comparable between these two approaches (Supplementary Table 3).

### Allele-selective siRNA treatment of HD cell lines

Allele-selective siRNA treatment of patient-derived HD cell lines GM04022 and GM02147 from the Coriell Institute was performed to demonstrate the assay’s ability to quantify knockdown of individual HTT alleles in a therapeutic context. To test allele-specific HTT transcript lowering, allele-selective siRNAs (targeting either the A or C allele at SNP rs363125) and a non-specific negative control siRNA were used at two doses (10 nM and 100 nM) (Fig. 3a). The GM04022 cell-line is heterozygous (A/C) for the siRNA target SNP, with the A-allele being in phase (i.e. in the same haplotype) as the wtHTT (shorter CAG repeats) and the C-allele being in phase with the mHTT (longer CAG repeats) (Fig. 3a). The GM02147 cell-line is homozygous C for the siRNA target SNP, i.e. both the wtHTT and mHTT alleles are in phase with the C-allele (Fig. 3a). Therefore, given the genotypic differences between the two treated cell lines, we expected different HTT allele-specific lowering patterns when treating with A- or C-selective siRNAs (Fig. 3a). Results obtained with our assay showed that the proportion of the mutant HTT allele changed as expected depending on the treatment with the allele-selective siRNAs and the genotype of cell line being treated (Fig. 3b and c). Namely, the mHTT:wtHTT ratio was only affected within the heterozygous cell-line GM04022 (Fig. 3b and c). The dose of the siRNA treatment (10 nM vs 100 nM) had very little or no effect on allele-selective HTT mRNA lowering. These results confirm the precision of our assay in distinguishing allele-selective lowering following the treatment.

**Fig. 3.**
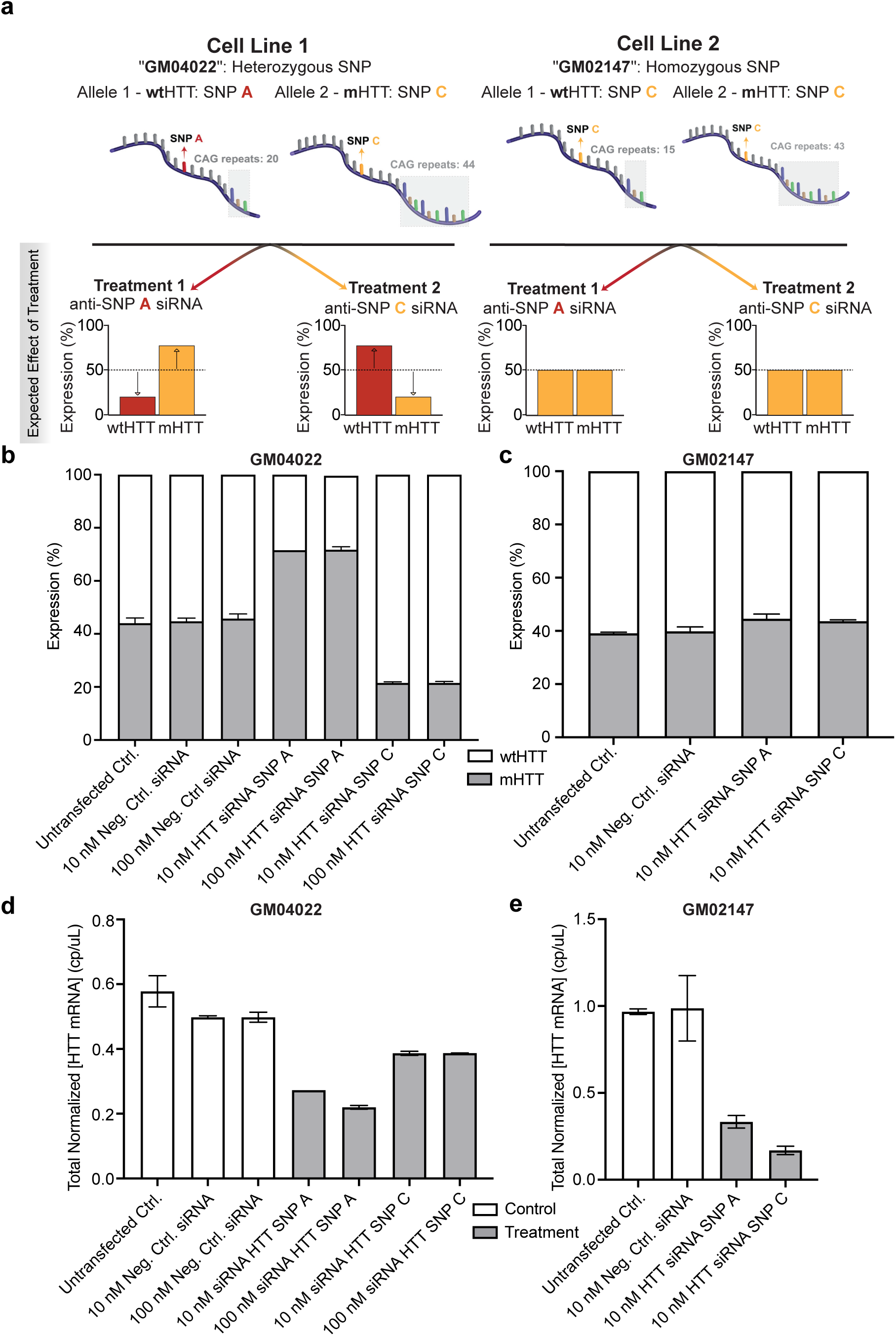
HTT mRNA lowering after treatment with allele-selective siRNAs. **a)** Experimental setup and expected outcome of treating two different HD Coriell cell-lines (GM04022 and GM021477) with two different allele-selective siRNAs. **b-c)** Proportion (%) of mHTT and wtHTT expression in GM04022 (A/C genotype) (**b**) and GM02147 (C/C genotype) (**c**) cell-lines after treatment with allele-selective and negative control siRNAs in different doses and two biological replicates. In the heterozygous (A/C) Cell-line 1, A-selective siRNA increases the mHTT proportion by lowering wtHTT expression, while C-selective siRNA decreases the mHTT proportion by lowering mHTT expression. In the homozygous (C/C) Cell-line 2, the mHTT:wtHTT proportion remains unchanged with A-selective siRNA treatment, as there is no treatment effect on either allele. Similarly, the mHTT:wtHTT proportion stays the same with the C-selective siRNA treatment, as it lowers the expression of both alleles. **d-e)** Total HTT mRNA lowering in GM04022 **(d)** and GM02147 **(e)** by analysing the both clusters together and normalised by the GUSB housekeeping gene and the expression levels of the untransfected sample. The 10 nM SNP A result for GM04022 did not have a replicate.

Additionally, using the same data, we calculated overall HTT expression levels by normalizing the total detected HTT mRNA values with the GUSB housekeeping gene mRNA levels measured in a separate reaction (Fig. 3d and e). We found that the siRNAs targeting A and C SNP alleles are allele-selective rather than completely allele-specific, and thereby we saw HTT lowering with both siRNAs, regardless of the genotype of the treated cell-line (Fig. 3d and e). However, the lowering effect was stronger when the siRNA used matched perfectly with the HTT mRNA target sequence (Fig. 3d and e). As expected, the negative control siRNA does not change the allele proportions or overall HTT levels (Fig. 3d and e).

## Discussion

Here we have demonstrated that the accurate detection of allele-specific HTT mRNA lowering is possible with ddPCR in specific dNTP depleted conditions without a need for allele-specific or SNP-targeting primer/probe design. This assay has great potential to be used as a pharmacodynamic (PD) biomarker in clinical trials aiming for allele-selective HTT mRNA lowering. We started the assay optimization with the conditions described by Dodd et. al. (12) who showed preliminary data of one HD patient derived Coriell sample in non-optimised conditions. For example, a very low number of negative droplets in their experiment would prevent accurate concentration measurement according to the Poisson distribution; we made sure that more than half of the droplets were negative. We also demonstrated that simplifying the assay into 1-step RT-ddPCR instead of the 2-step RT-PCR originally described (12) did not alter the results. Added benefits of this approach are more straightforward and faster lab procedure, and reduced number of potentially error-prone steps, while not affecting the sensitivity or accuracy of the assay. Finally, we extended the experiment to include 13 Coriell fibroblast cell lines derived from Huntington’s disease patients and family members, including siRNA experiments to show a robust quantification of allele-selective lowering.

In addition to the optimized lab protocol, we have developed an algorithm for automated thresholding between the wild-type and mutant allele signals, followed by allele-specific concentration measurements based on poisson distribution. This has two advantages: first, any manual and potentially subjective manipulation becomes unnecessary, enabling fully objective allele-specific concentration calculations. Second, the poisson distribution-based concentration calculation is performed for the two different alleles in an identical way, because everything other than the target cluster is considered as a negative cluster.

When the assay is used for detecting changing proportions of mHTT and wtHTT expression, we do not need to normalize the results using a housekeeping gene expression as a reference value. However, if the goal is to detect total HTT lowering, the use of a housekeeping gene for normalization is recommended despite the absolute quantification nature of ddPCR. Due to the assay design (depleted dNTP conditions), multiplexing with a housekeeping gene is not possible. Therefore, if a housekeeping gene result is required, it has to be included as a separate ddPCR assay. While potential pipetting bias may slightly confound the results in this scenario, running a housekeeping gene assay in parallel can mitigate other biases, such as cDNA conversion. Although with this assay design the amplitude of the fluorescence signal is dependent on the amplicon length, it will not have an influence on the total HTT mRNA expression level. This is because the concentration readout in ddPCR is based on the number of positive droplets and not on the fluorescence signal intensity. Nevertheless, even though the main use case of the assay is the allele-specific mRNA expression level detection, its ability to also detect overall HTT lowering is beneficial in order to identify any potential non-allele specific treatment effects.

The phenomenon of HTT allelic-imbalance (i.e. wtHTT and mHTT alleles being expressed at varying levels) described previously in (13) is also clear in our data sets. The proportion of mHTT mRNA expression in heterozygous Coriell cell lines (n=8) analyzed in this study varied from 24.6% to 42.1% (mean 34.3% with a negative correlation with CAG repeat length). This is in line with what was described (31–38% mHTT proportion) with experiments also performed on HD patient derived cell lines (13). However, in contrast to the in vitro findings in cell lines, a study of post-mortem HD patient brains suggests higher mHTT than wtHTT mRNA levels in vivo, particularly in the HD critical brain regions such as striatum and cortex (21). Shin *et al*. (22) reported, contrary to our findings, that there is no correlation between the HTT expression levels and the CAG repeat size in lymphoblastoid cell-lines. A possible reason for differential mHTT and wtHTT mRNA expression level detection could be that the reverse transcription efficiency is influenced by mRNA secondary structures, leading to lower recovery of the longer mHTT mRNA. However, this potential difference in baseline detection does not compromise the assay’s utility, as its primary purpose is to detect the pharmacodynamic effects of a drug, specifically the change from baseline after treatment.

It is important to acknowledge that the ddPCR assay, while precise, has limitations in distinguishing very small differences in CAG repeat length. Our experiments indicate that with the right bandwidth (*h*) value the assay and its computational pipeline can reliably differentiate alleles with a difference of at least 10% in repeat length (as demonstrated by the Coriell sample GM04791 with 18/20 repeat length units). This bandwidth tuning allows the user to define the assay’s sensitivity allowing even extremely small droplet clusters to be detected. For the purposes of HTT transcript detection with ddPCR however we found that a bandwidth value of between 90 and 100 was ideal to prevent background droplets being included as real clusters. Nevertheless, CAG repeat length differences such as those observed in the mHTT homozygous alleles of GM13511 with 45 and 47 CAG repeat length units (a 4.3% difference in length), may fall within the assay’s margin of error and are no longer visually or computationally distinguishable. However, it’s crucial to note that in the context of HD, this limitation has minimal clinical impact. Full-penetrance mutations typically involve CAG repeat lengths of 40 or more, while reduced-penetrance mutations range from 36 to 39 repeats (8). Thus, virtually all heterozygous HD patients present with an allele length difference exceeding 10%, ensuring that the assay’s resolution is sufficient for accurately distinguishing between mutant and wild-type alleles in clinical applications.

The principle of this ddPCR assay, utilizing dNTP depleted conditions to differentiate between repeat lengths, could potentially be extended to other repeat expansion diseases beyond Huntington’s Disease. For example, different types of Spinocerebellar Ataxia (SCA), which are also caused by trinucleotide repeat expansions (23–25), may be well-suited for this approach. While this highlights the potential of the broader applicability of the assay principle for studying various neurological disorders characterized by repeat expansions it’s important to note that this method may not be suitable for repeat expansion disorders with extremely long repeat lengths. For instance, Myotonic Dystrophy Type 1 (DM1), which may involve thousands of CTG repeats (26,27), might pose challenges to amplify the longer repeat efficiently.

Another consideration is the somatic instability of the CAG repeat region, particularly in the mutant HTT allele (2). While somatic expansion is a significant factor in brain tissues and potentially in the CSF, it is generally less pronounced in peripheral tissues (2,28,29). Importantly, the wild-type HTT allele is not expected to undergo significant somatic expansion, providing a relatively stable reference point for our ddPCR assay (30,31). Therefore, even in the presence of some somatic instability in the mutant allele, we can still reliably differentiate between the wild-type and mutant HTT transcripts. However, it is conceivable that in cases of extreme somatic expansion, the mutant allele may become so long that it is no longer amplified. In such instances, our assay would reflect a decrease in the detected expression of those extremely expanded mutant molecules. This potential effect, while unlikely to significantly confound the overall quantification of allele-specific HTT mRNA expression, should be considered when interpreting results from samples with known or suspected high levels of somatic instability. Therefore, it is unlikely that somatic expansion significantly impacts the accuracy of our assay for quantifying allele-specific HTT mRNA expression.

Taken together, our assay together with its automated analysis pipeline fills an important gap in the biomarker repertoire for HD allowing allele-specific detection of HTT mRNA expression levels. Other methods described in the literature rely on SNPs in the HTT locus that are in LD with the CAG repeat in exon 1 (13,21,22). As mentioned, this approach is not ideal, as the LD structure defining the HTT haplotypes is population-specific. Moreover, LD between the CAG repeat and SNPs in the HTT locus is incomplete in any population, meaning that some patients are homozygous for the assay target SNP, which would prevent the allele-specific mHTT mRNA detection. Therefore, unless haplotype information is available for every patient sample, SNP-based mHTT mRNA detection assays cannot be considered sufficiently reliable for clinical applications.

## Conclusions

We have demonstrated that the accurate detection and quantification of allele-specific HTT mRNA ratios and their lowering is possible with ddPCR under specific dNTP depleted conditions, without the need for allele-specific primer/probe design. Our method allows for the precise detection of relative ratios between mHTT and wtHTT transcripts, as well as the quantification of total HTT mRNA levels when normalized using an internal housekeeping gene. Furthermore, we have developed an analysis pipeline to process dNTP depleted ddPCR results in an unbiased and automatic manner. This unique assay principle and its computational pipeline may serve a critical need for robust and quantitative biomarker measurements in support of the development of allele-selective HTT lowering therapies in HD.

## Data Availability

All data produced in the present study are available upon reasonable request to the authors

## List of abbreviations

ASO: Antisense Oligonucleotide
cDNA: Complementary DNA
ddPCR: Droplet Digital PCR
DM1: Myotonic Dystrophy Type 1
dNTP: Deoxynucleotide Triphosphate
GUSB: Glucuronidase Beta (housekeeping gene)
HD: Huntington’s Disease
HTT: Huntingtin Gene
KDE: Kernel Density Estimation
LD: Linkage Disequilibrium
LOF: Local Outlier Factor
mHTT: mutant huntingtin
wtHTT: wild-type huntingti
mRNA: Messenger RNA
NIGMS: National Institute of General Medical Sciences
PCR: Polymerase Chain Reaction
PD: Pharmacodynamic
qPCR: Quantitative
PCR RNA: Ribonucleic Acid
RT-ddPCR: Reverse Transcription Droplet Digital
PCR SCA: Spinocerebellar Ataxia
siRNA: Small Interfering RNA
SNP: Single Nucleotide Polymorphism
1D: One Dimensional
2D: Two Dimensional

## Declarations

## Ethics approval and consent to participate

All human-derived cell lines and DNA samples used in this study were obtained from NIGMS Human Genetic Cell Repository at the Coriell Institute for Medical Research. All samples in the Coriell repositories are collected with appropriate informed consent from the donors

## Consent for publication

All authors have consented for the publication of the manuscript

## Availability of data and materials

All data generated or analysed during this study are included in this published article and its supplementary information files

## Authors’ contributions

E.G. Performed the data-analyses with the help of D.R., E.G. prepared the figures and tables, E.G., D.R., M.T. and A.R. contributed to the early drafting of the manuscript, M.T. performed the RNA extractions and ddPCR experiments, J.M. and A.M. performed the siRNA experiments, N.M., D.J.H and M.S. gave expert input on data interpretation and manuscript writing, A.R. conducted the study design, supervised the study and finalised the manuscript with the input from all co-workers. All authors read and approved the final manuscript.

## Acknowledgements

The authors would like to thank all individuals who contributed to this work. All major contributions are reflected in the authorship list. We would also like to thank the Coriell Institute for Medical Research for providing the human cell lines used in this study. The following samples were obtained from the Coriell Institute for Medical Research: GM04282, GM04691, GM04724, GM04791, GM04819, GM04855, GM04859, GM04869, GM13504, GM13511, GM21757, GM04022, and GM02147.

