## Supplementary Tables for "Droplet digital PCR assay to analyze allele-specific mRNA expression on HTT repeat expansion locus"

| Coriell ID | mHTT Concentration (cp/μL) | wtHTT Concentration (cp/μL) | Diagnosis | CAG Repeat Length (Coriell) | CAG Repeat Length (Seq) | mHTT % |
|---|---|---|---|---|---|---|
| GM04282 | 127.83 | 391.42 | HD | NA | 73/18 | 24.6 |
| GM04691 | 95.36 | 153.01 | HD | NA | 55/16 | 38.4 |
| GM04724 | 110.58 | 303.27 | HD | NA | 69/16 | 26.7 |
| GM04791 | NA | 364.40 | WT | NA | 20/18 | 0.0 |
| GM04819 | 141.99 | 202.65 | HD | NA | 45/18 | 41.2 |
| GM04855 | 109.48 | 182.67 | HD | NA | 49/21 | 37.5 |
| GM04859 | NA | 223.19 | WT | NA | 20/20 | 0.0 |
| GM04869 | 89.07 | 122.64 | HD | NA | 48/16 | 42.1 |
| GM13504 | 289.95 | 582.01 | HD | 46/16 | 47/17 | 33.3 |
| GM13511 | 397.58 | NA | HD | 47/45 | NA | 100.0 |
| GM21757 | 169.22 | 379.32 | HD | 66/16 | NA | 30.8 |
| GM04022* | NA | NA | HD | NA | 20/44 | NA |
| GM02147* | NA | NA | HD | NA | 15/43 | NA |

**Supplementary Table 1**. Coriell samples used for the assay optimisation and proof of concept experiments. Detected allele-specific mRNA expression of 11 of the 13 different Coriell samples. Samples shown with (*) were used for siRNA experiments only.

| Input mHTT (%) | mHTT Concentration (cp/μL) | wtHTT Concentration (cp/μL) | % mHTT Detected | Average % mHTT Detected (st. dev) |
|---|---|---|---|---|
| **100** | 170.16 | 0.00 | 100.00 | 100.00 (±0) |
| **100** | 150.01 | 0.00 | 100.00 | |
| **100** | 165.87 | 0.00 | 100.00 | |
| **80** | 134.96 | 30.00 | 81.81 | 83.82 (± 1.74) |
| **80** | 140.10 | 25.05 | 84.83 | |
| **80** | 138.64 | 24.83 | 84.81 | |
| **60** | 101.21 | 67.48 | 60.00 | 61.15 (±1.10) |
| **60** | 101.02 | 61.44 | 62.18 | |
| **60** | 98.31 | 62.13 | 61.28 | |
| **50** | 85.12 | 81.24 | 51.17 | 50.74 (±0.37) |
| **50** | 81.14 | 79.24 | 50.59 | |
| **50** | 83.13 | 81.56 | 50.48 | |
| **40** | 65.17 | 105.79 | 38.12 | 38.38 (±0.73) |
| **40** | 65.46 | 101.48 | 39.21 | |
| **40** | 63.22 | 103.94 | 37.82 | |
| **20** | 34.94 | 144.09 | 19.52 | 18.53 (±1.14) |
| **20** | 32.58 | 140.83 | 18.79 | |
| **20** | 29.46 | 141.02 | 17.28 | |
| **0** | 0.00 | 187.25 | 0.00 | 0.00 (±0) |
| **0** | 0.00 | 172.31 | 0.00 | |
| **0** | 0.00 | 167.86 | 0.00 | |
| **NTC** | 0.00 | 0.00 | 0.00 | 0.00 (±0) |
| **NTC** | 0.00 | 0.00 | 0.00 | |
| **NTC** | 0.00 | 0.00 | 0.00 | |

**Supplementary Table 2**. Simulation of the allele-specific lowering by mixing cDNA from mHTT (GM13511; 45/47 CAG repeats) and wtHTT (GM04859; 20/20 CAG repeats) homozygotes in known proportions. Observed concentrations for mHTT and wtHTT clusters and observed proportion of detected mHTT are shown in 3 replicates.

| Sample Name (HTT genotype) | 1-step RT-ddPCR | | 2 step RT-ddPCR | |
|---|---|---|---|---|
| | Proportion of WT HTT conc. (%) | Proportion of Mutant HTT conc. (%) | Proportion of WT HTT conc. (%) | Proportion of Mutant HTT conc. (%) |
| **GM04282 (77/18)** | 75.4 | 24.6 | 75.4 | 24.6 |
| **GM04691 (57/16)** | 64.3 | 35.7 | 61.6 | 38.4 |
| **GM04724 (71/16)** | 72.7 | 27.3 | 73.3 | 26.7 |
| **GM04791 (18/20)** | 100.0 | 0.0 | 100.0 | 0.0 |
| **GM04819 (45/18)** | 58.8 | 41.2 | 58.8 | 41.2 |
| **GM04855 (48/21)** | 64.8 | 35.2 | 62.5 | 37.5 |
| **GM04859 (21/17)** | 100.0 | 0.0 | 100 | 0.0 |
| **GM04869 (48/16)** | 63.8 | 36.2 | 57.9 | 42.1 |

**Supplementary Table 3.** Comparison of 1-step RT-ddPCR vs 2-step RT-ddPCR approach on 8 Coriell cell lines.
