## Supplementary Figures for "Droplet digital PCR assay to analyze allele-specific mRNA expression on HTT repeat expansion locus"

### Outliers Removal Example

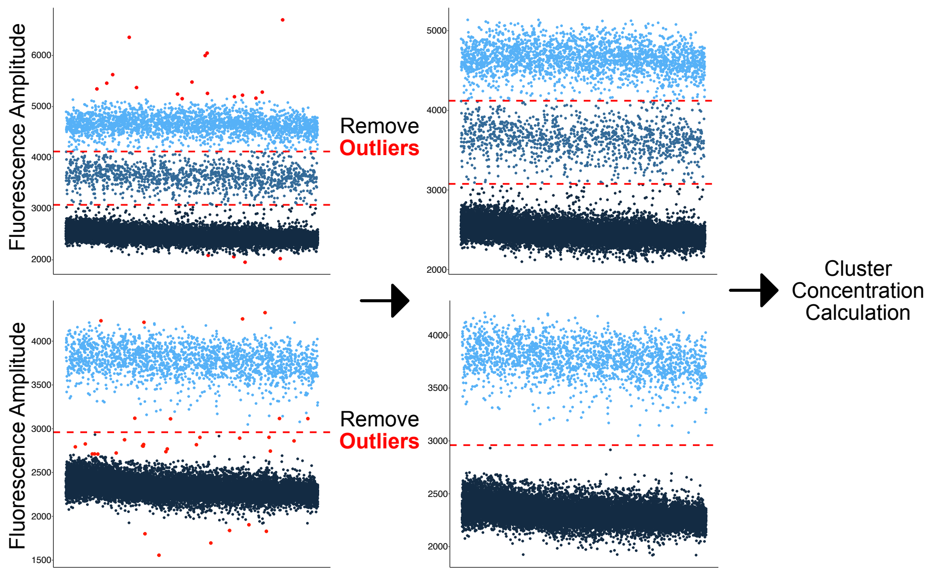

**Supplementary Fig. 1. Outlier detection and removal process before cluster concentration calculation.** Outliers are identified using the local outlier factor (LOF) algorithm. Identified outliers are shown in red in the example above.

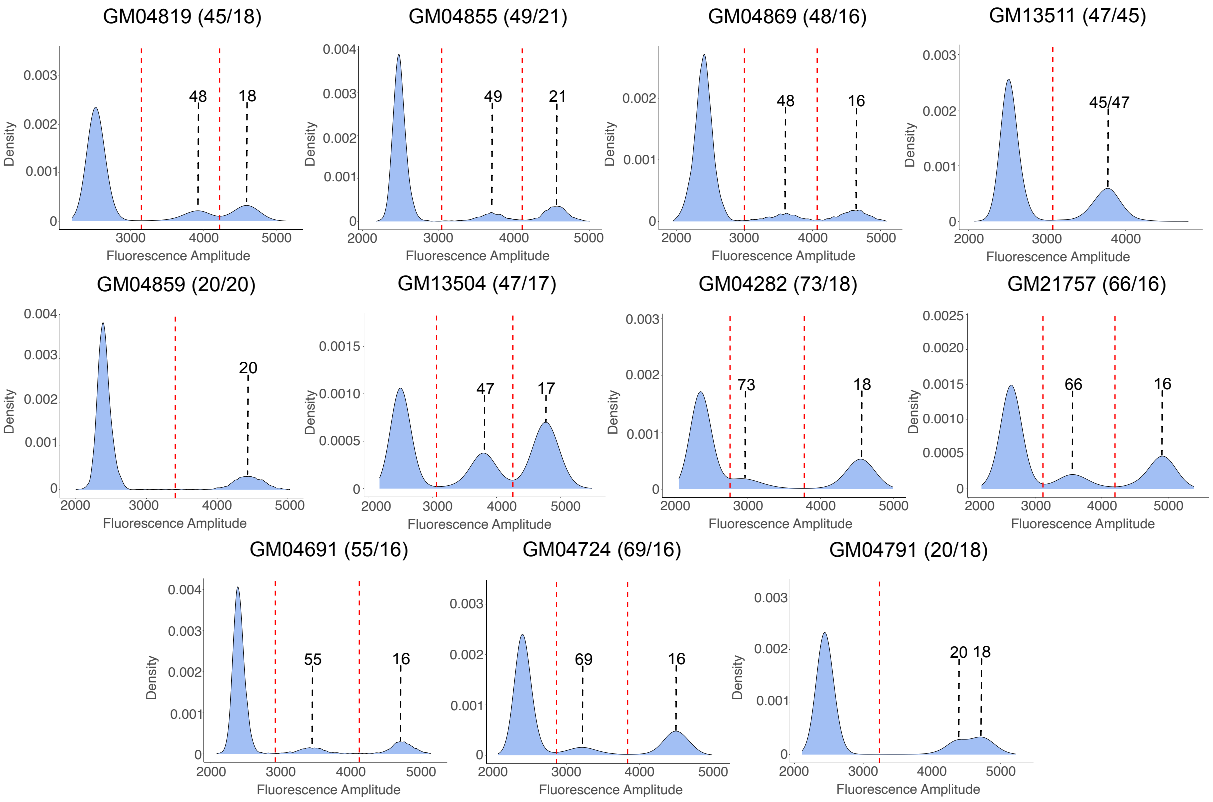

GM04819 (45/18)
48
18
GM04855 (49/21)
49
21
GM04869 (48/16)
48
16
GM13511 (47/45)
45/47
GM04859 (20/20)
20
GM13504 (47/17)
47
17
GM04282 (73/18)
73
18
GM21757 (66/16)
66
16
GM04691 (55/16)
55
16
GM04724 (69/16)
69
16
GM04791 (20/18)
20 18
Density
Fluorescence Amplitude

**Supplementary Fig, 2. Allelic separation of HTT CAG repeat region in 11 Coriell cell lines with non-diluted cDNA starting material**. The positive droplet clusters are marked based on the known HTT CAG repeat length (determined by sequencing).

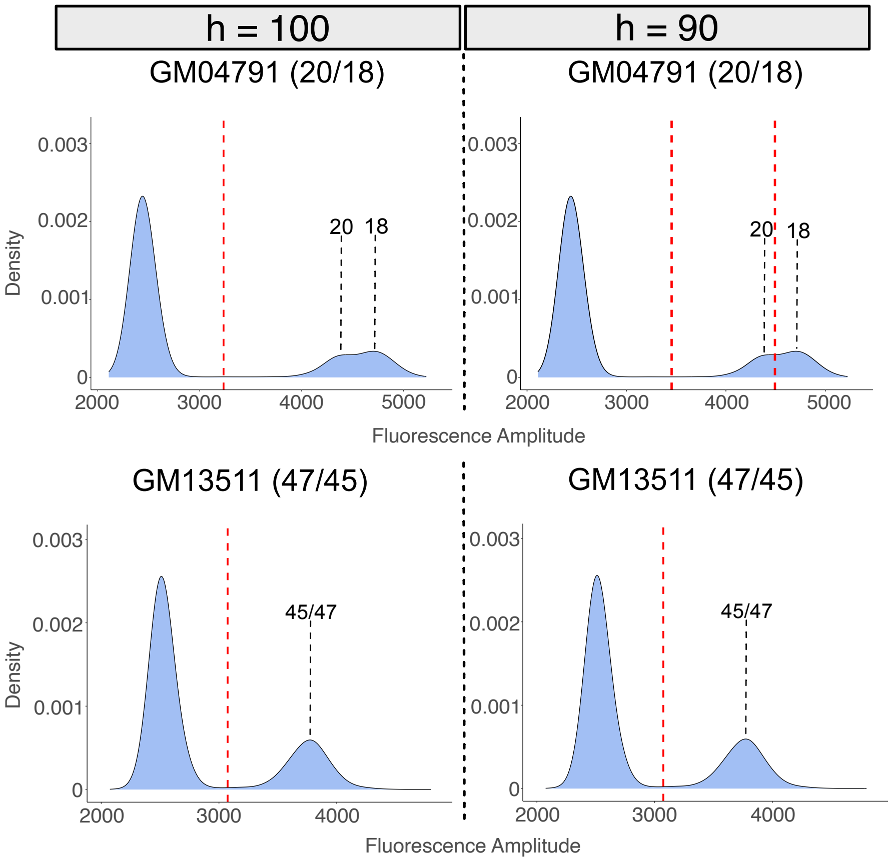
h = 100
h = 90
GM04791 (20/18)
GM04791 (20/18)
Density
0.003
0.002
0.001
0
20
18
2000
3000
4000
5000
Fluorescence Amplitude
GM13511 (47/45)
GM13511 (47/45)
45/47
45/47
Fluorescence Amplitude

**Supplementary figure 3. Lowering the bandwidth value *h* increases the sensitivity of the automatic analysis pipeline allowing the differentiation of very closely overlapping clusters of GM04791.** The increased sensitivity does not come at the cost of decreased specificity as the pipeline does separate otherwise unique clusters such as the GM13511 peak.

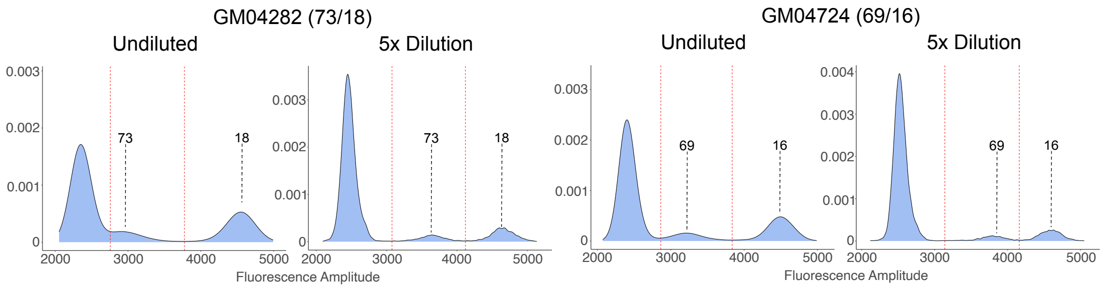
GM04282 (73/18)
Undiluted
5x Dilution
0.003
0.002
0.001
0
73
18
2000
3000
4000
5000
Fluorescence Amplitude
GM04724 (69/16)
Undiluted
5x Dilution
0.004
69
16

**Supplementary Fig. 4. Examples of 2 instances where 5x dilution of input cDNA yielded improved separation of high CAG repeat droplet clusters from negative clusters.**
